# Middle-aged individuals may be in a perpetual state of H3N2 influenza virus susceptibility

**DOI:** 10.1101/2020.01.09.20017038

**Authors:** Sigrid Gouma, Kangchon Kim, Madison Weirick, Megan E. Gumina, Angela Branche, David J. Topham, Emily T. Martin, Arnold S. Monto, Sarah Cobey, Scott E. Hensley

**Author notes:** corresponding author: 402 Johnson Pavilion, 3610 Hamilton Walk, Philadelphia, PA 19104.

## Abstract

Most humans are infected with influenza viruses by 3-4 years of age (1) and have high antibody titers against viral strains encountered early in life (2). Early childhood influenza exposures can leave lifelong ‘immunological imprints’ that affect how an individual responds to antigenically distinct viral strains later in life (3,4). H3N2 influenza viruses began circulating in humans in 1968 and have evolved substantially over the past 51 years (5). Therefore, an individual’s birth year largely predicts which specific type of H3N2 virus they first encountered in childhood. Here, we completed a large serological survey to elucidate the specificity of antibodies against contemporary H3N2 viruses in differently aged individuals who were likely primed with different H3N2 strains in childhood. We found that most humans who were first infected in childhood with H3N2 viral strains from the 1960s and 1970s possess non-neutralizing antibodies against contemporary 3c2.A H3N2 viruses. Most importantly, we found that 3c2.A H3N2 virus infections boost non-neutralizing H3N2 antibodies in middle-aged individuals, potentially leaving many of them in a perpetual state of 3c2.A H3N2 viral susceptibility.

---

An antigenically drifted descendant of the original 1968 H3N2 strain, designated as clade 3c2.A H3N2 viruses, emerged during the 2014-2015 influenza season (6) and continues to circulate at high levels across most parts of the world (7). 3c2.A H3N2 viruses are unique because they possess a key glycosylation site that shields HA antigenic site B, which is a major target of neutralizing antibodies (8). During the 2017-2018 season, 3c2.A H3N2 viruses caused a lot of hospitalizations and deaths in the United States (9). Influenza activity is often first detected in children within the community; however, during the severe 2017-2018 influenza season, influenza activity in middle-aged and older adults peaked earlier than in children and young adults (9). It is surprising that 3c2.A H3N2 viruses dominated the 2017-2018 season in the United States since these viruses also dominated the 2014-2015 and 2016-2017 United States influenza seasons, and therefore many people likely had immunity against these viruses. It is unknown if 3c2.A viruses were able to circulate widely in 3c2.A H3N2-exposed populations during the 2017-2018 season by acquiring HA and NA substitutions that facilitate antibody escape. Further, it is unknown why these viruses exhibited unusual age-related patterns of circulation during that season and whether or not this was a consequence of different H3N2 exposure histories in different aged individuals.

To determine if HA and/or NA substitutions in 3c2.A H3N2 viruses contributed to the atypical 2017-2018 influenza season in the United States, we completed a large serological survey using samples collected from differently aged individuals in the summer months prior to the 2017-2018 influenza season. We tested sera from individuals with different birth years so that we would be able to identify antibody signatures that were associated with distinct H3N2 childhood immune imprinting. In total, we tested serum samples collected from 140 children (age 1-17) at the Children’s Hospital of Philadelphia and 212 adults (age 18-90) at the Hospital of the University of Pennsylvania (Extended Data Table 1).

First, we used ELISA to measure antibody binding to recombinant HA proteins from a 3c2.A virus from 2014 (herein referred to as 3c2.A) and from a 3c2.A virus from 2018 (herein referred to as 3c2.A2). The HA of 3c2.A2 H3N2 viruses from 2018 possesses 3 amino acid substitutions (T131K, R142K, and R261Q) relative to the HA of 3c2.A H3N2 viruses from 2014 (10). Most individuals 4 years of age and older possessed 3c2.A and 3c2.A2 HA ELISA-reactive antibodies (Fig. 1a and d). Only 38 of 352 serum samples had no detectable HA reactive antibodies. Thirty-five of the 38 donors with no detectable HA-reactive antibodies were under 4 years of age, and it is likely that these individuals were not previously exposed to H3N2 virus. Importantly, we found no major differences in mean serum antibody binding to 3c2.A HA compared to 3c2.A2 HA (Fig. 1g).

**Figure 1.**
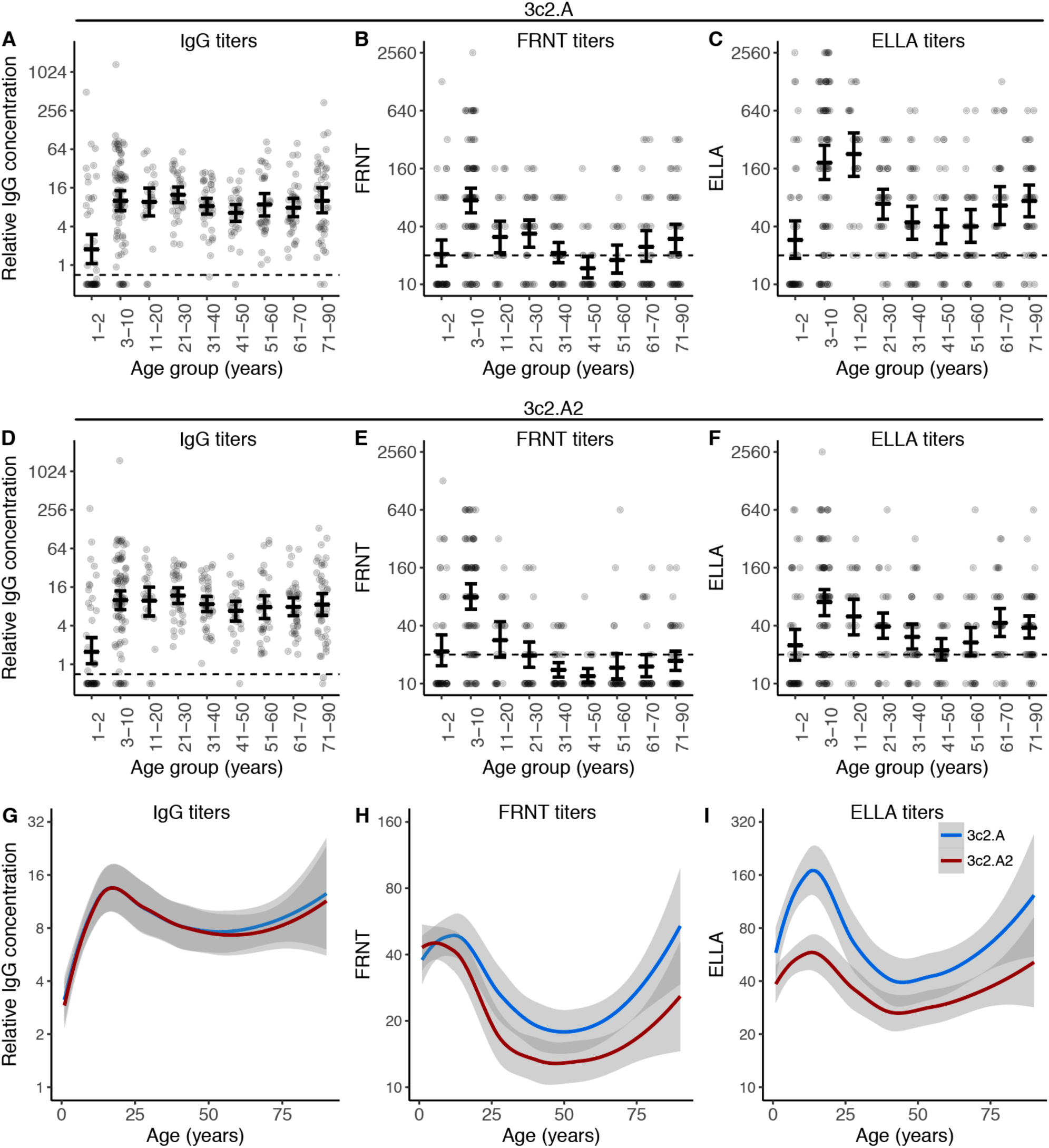
Antibody titers to contemporary A(H3N2) viruses in serum collected prior to the 2017-2018 season. ELISAs were completed to quantify the levels of 3c2.A (A) and 3c2.A2 (D) HA-specific IgG in serum from different aged humans. FRNTs were completed to quantify the levels of 3c2.A (B) and 3c2.A2 (E) neutralizing antibodies in serum from different aged humans. ELLAs were completed with viruses that had 3c2.A NA (C) and 3c2.A2 NA (F) to quantify NA-specific inhibitory antibodies in serum from different aged humans. For A-F, the geometric mean titers are shown as cross bars and 95% confidence intervals as error bars. (G-H) Data from panels A-F are summarized. Lines (blue=3c2.A; red=3c2.A2) are Loess curves showing smoothed geometric mean titers with the 95% confidence intervals shaded. Dashed lines in panels A-F represent the limit of detection for each assay.

Our ELISAs are designed to detect all HA-binding antibodies, and these assays cannot distinguish between neutralizing and non-neutralizing antibodies. Therefore, we next completed neutralization assays using 3c2.A and 3c2.A2 viruses. These assays primarily measure neutralizing HA antibodies and do not efficiently detect non-neutralizing HA or most NA antibodies. Contrary to what we found in ELISAs, most individuals did not possess high levels of neutralizing antibodies against either 3c2.A or 3c2.A2 viruses (Fig. 1b). Sera from children aged 3-10 years old possessed the highest levels of neutralizing antibodies against these viruses (p<0.0007 when compared to all other age groups for both 3c2.A and 3c2.A2 titers, Extended Data Table 2), whereas most middle-aged adults did not have detectable neutralizing antibody titers. Among adults, neutralizing antibody titers were lowest in individuals ∼50 years of age. This is notable since 50 year olds were born in 1967, one year before H3N2 viruses began circulating in humans. Neutralizing antibody titers against 3c2.A2 virus were lower compared to titers against 3c2.A virus in serum from some individuals, but only 37 individuals (11%) had a ≥4-fold decrease in neutralizing antibody titer to this virus (Fig. 1h and Extended Data Fig. 1). These data suggest that 2017-2018 H3N2 viruses did not possess significant HA antigenic changes relative to 2014-2015 H3N2 viruses for most humans; however a large fraction of the adult population possessed non-neutralizing antibodies that could bind to the HAs of these viruses but could not prevent virus infection.

We next completed experiments to measure serum antibody binding to NA. Antibodies against NA typically do not neutralize virus infection (11), so we completed Enzyme-Linked Lectin Assays (ELLAs) that measure antibodies that block NA cleavage of sugars on fetuin proteins (12). Similar to HA neutralizing antibody titers, NA-specific antibody titers were highest in younger individuals and lowest in middle-aged adults (Fig. 1c). Consistent with a recent report (13), we found that some individuals possessed antibodies that reacted to the NA of 3c2.A viruses more efficiently compared to the NA of 3c2.A2 viruses (Fig. 2c, f and i). The NA of 3c2.A2 viruses possesses a new glycosylation site at residue 245 (enabled by S245N+S247T substitutions relative to the NA of 3c2.A NA) that alters antibody binding (13). Interestingly, antibodies that were able to antigenically distinguish the 3c2.A and 3c2.A2 NAs were more prevalent in individuals younger than 25 years of age (Fig. 1c, f and i).

**Figure 2.**
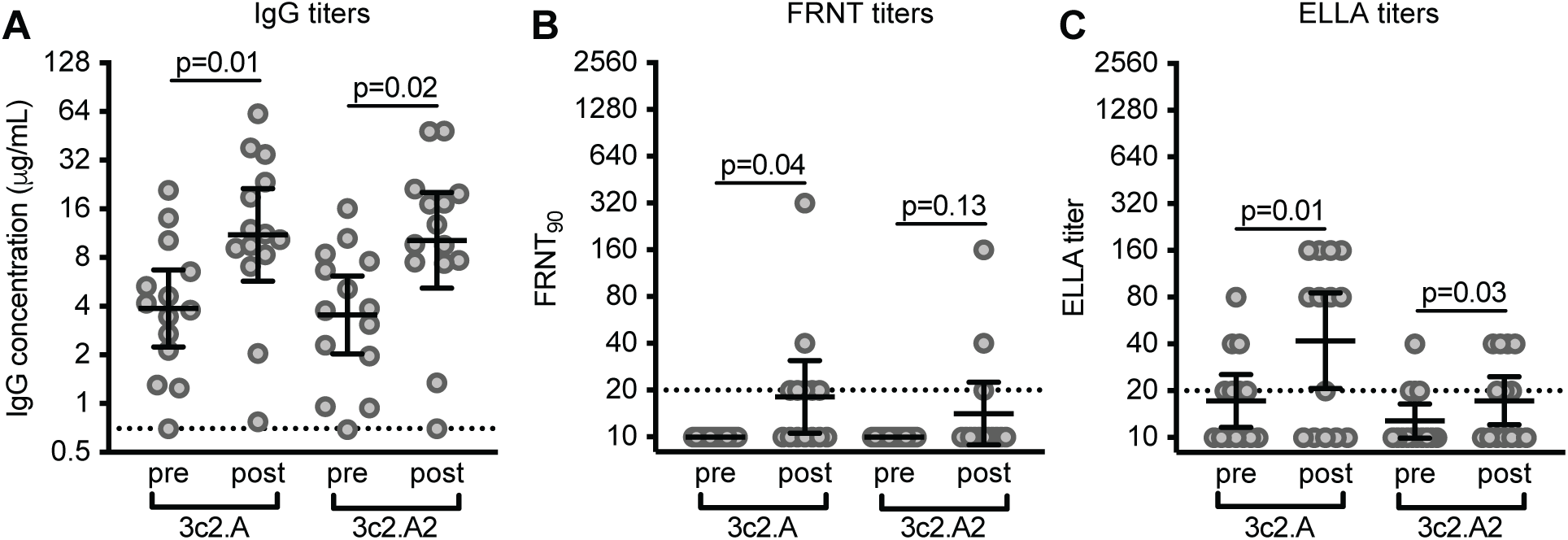
Antibody responses in adults before and after laboratory-confirmed 3c2.A H3N2 infection during the 2014-2015 influenza season. Serum was collected from 14 adults before and after laboratory-confirmed 3c2.A H3N2 infection and we quantified (A) HA IgG binding antibodies by ELISA, (B) neutralizing antibodies by FRNTs, and (C) NA-specific antibodies by ELLAs. Dashed lines represent the limit of detection for each assay. Lines represent the geometric mean titers with 95% confidence intervals. Paired t tests were completed on log_2_-transformed data, and p values are indicated.

It is unclear why many adults born ∼1968 possessed low levels of 3c2.A and 3c2.A2 HA neutralizing antibodies and NA antibodies prior to the 2017-2018 influenza season. It is possible, although unlikely, that 3c2.A viruses did not infect this age group during the previous 2014-2015 and 2016-2017 influenza seasons. An alternative hypothesis is that individuals born ∼1968 were infected as much as other adults during the previous 3c2.A H3N2-dominated seasons but failed to mount neutralizing antibody responses that could confer protection during the 2017-2018 season. To address this possibility, we measured HA and NA antibodies in 14 middle-aged adults (years of birth ranging from 1963-1979) before and after laboratory-confirmed 3c2.A H3N2 influenza virus infection during the 2014-2015 season. These samples were collected as part of the Household Influenza Vaccine Effectiveness (HIVE) study in Ann Arbor, Michigan (14). Consistent with the hypothesis that 3c2.A H3N2 viruses elicit a non-neutralizing antibody response in middle-aged adults, we found that HA ELISA antibody titers increased in the serum of most individuals 4-8 months after infection (Fig. 2a), but neutralizing antibody titers remained low in the same individuals (Fig. 2b). 3c2.A NA inhibitory antibodies increased in the serum of most infected individuals and these antibodies reacted less well to the antigenically drifted 3c2.A2 NA from 2018 (Fig. 2c). We found similar results when we analyzed serum samples from a separate smaller cohort of individuals (n=6) collected 28 days post-infection in Rochester, New York during the 2016-2017 season (Extended Data Fig. 2).

The HAs of 1968 H3N2 viruses and contemporary 3c2.A H3N2 viruses are dissimilar and most epitopes conserved between these HAs are non-neutralizing (Fig. 3a). We hypothesize that individuals ‘imprinted’ with H3N2 viruses in childhood in the 1960s and 1970s produce mostly non-neutralizing HA antibodies against 3c2.A viruses because most of the 3c2.A HA memory B cells in these individuals are those that target non-neutralizing epitopes. We further hypothesize that 3c2.A infections in younger individuals do not elicit the same non-neutralizing responses since younger individuals have been primed with viruses more closely related to 3c2.A (Fig. 3a) or they have not yet been primed with H3N2 at all. Since H3N2 viruses have evolved continuously since 1968, individuals born more recently were likely ‘imprinted’ with an H3N2 strain that shares some neutralizing HA epitopes with 3c2.A viruses compared to individuals born closer to 1968. To model this, we calculated the probability of being ‘imprinted’ with H3N2 for differently aged individuals, taking into account the similarity of the HAs of the ‘imprinting’ H3N2 viral strain with the HA of 3c2.A viruses (Fig. 3b). Similarity between the HA of the ‘imprinting’ H3N2 strain and the HA of 3c2.A viruses was determined based on amino acid sequences (see Methods). We compared our model predictions with 3c2.A H3N2 neutralizing antibody titers in differently aged individuals (Fig. 3b). Most older individuals were not likely imprinted with H3N2 viruses, since these viruses did not start circulating until 1968. Antibody titers against 3c2.A viruses in these older individuals were at similar levels compared to young individuals. Conversely, most individuals born after 1968 were likely imprinted with an H3N2 virus, and this imprinting probability varies for each year of birth after 1977 since H1N1 viruses dominated some influenza seasons after 1977. In our model, being imprinted with an H3N2 virus did not correlate with neutralizing antibody titers to 3c2.A viruses (Extended Data Table 3). However, similarity between the HAs of the imprinting H3N2 viral strain and the 3c2.A H3N2 viral strain had a positive effect on neutralizing antibody titers (p<0.001, Extended Data Table 3). Younger individuals who were imprinted with an H3N2 virus with an HA that is more similar to the HA of 3c2.A H3N2 virus were more likely to possess neutralizing 3c2.A H3N2 antibodies compared to middle-aged adults who were imprinted with an H3N2 virus that has an HA that is more distant compared to the HA of 3c2.A H3N2 viruses. Taken together, our imprinting model suggests that the specific type of H3N2 virus that an individual is exposed to early in life can affect the specificity of antibody responses against contemporary 3c2.A H3N2 viruses.

**Figure 3.**
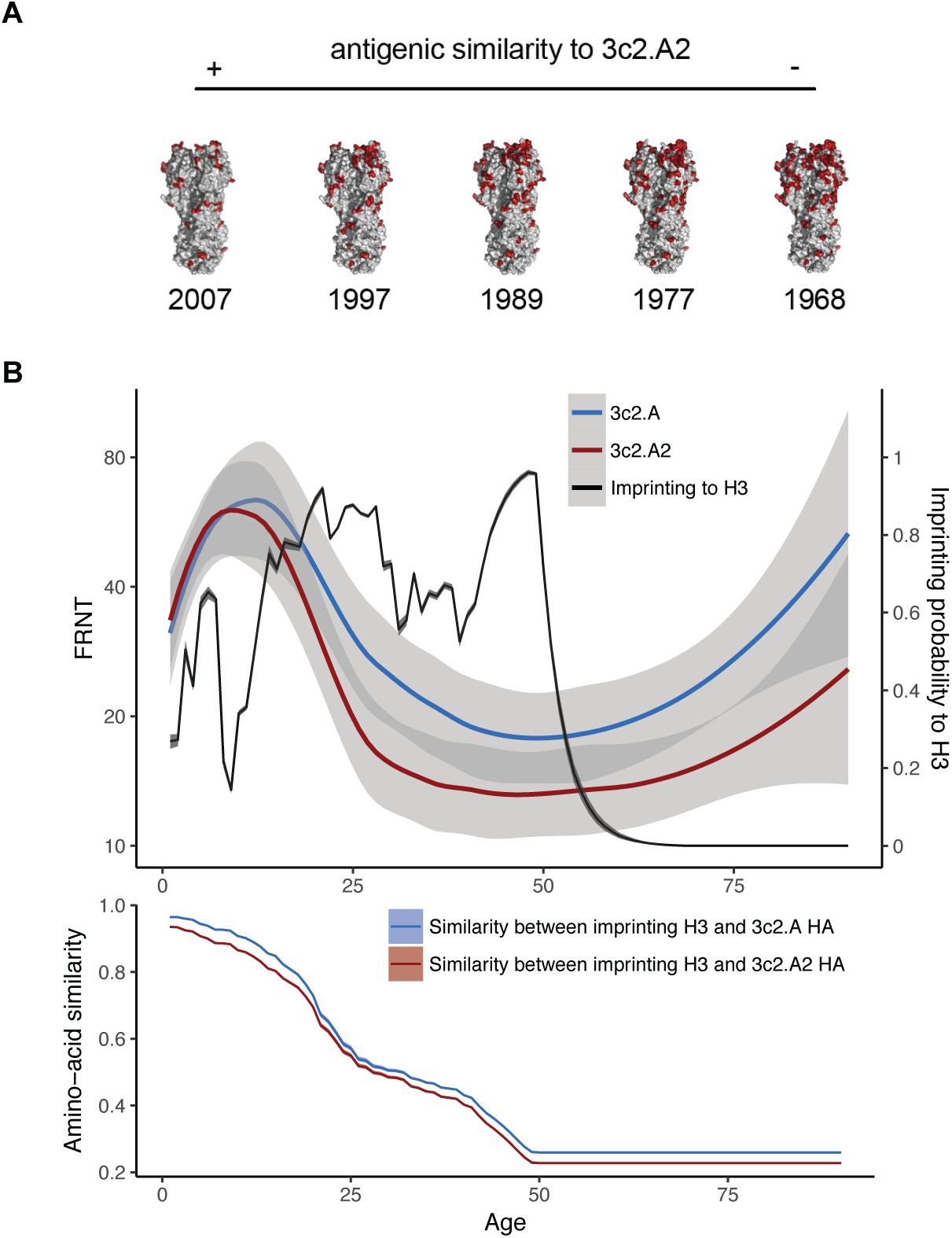
Early life H3N2 imprinting affects antibodies against 3c2.A viruses. (A) H3 has evolved substantially since 1968 and residue differences between the HA of different H3N2 strains and the 3c2.A2 strain are indicated in red. Accession numbers EF409245 (1968), CY113261 (1977), U97740 (1989), AJ311466 (1997) and EU199250 (2007) were aligned to accession number MH586372 (3c2.A2). (B) Top, mean FRNT titers to 3c2.A (blue) and 3c2.A2 (red) viruses are shown by Loess curves (span=0.6) with the 95% CIs shaded. The probability that an individual was imprinted with H3N2 (i.e. their first influenza A virus encounter was H3N2) is indicated with black line. Bottom, the antigenic similarity between the HA of an individual’s imprinting H3N2 strain and 3c2.A and 3c2.A2.

We propose that middle-aged individuals may be continuously susceptible to 3c2.A H3N2 infection since we found that these viruses preferentially elicit non-neutralizing HA antibody responses in this age group. While non-neutralizing HA and NA antibodies can stop viral spread and help resolve infections, these antibodies do not typically prevent infections. Although speculative, it is possible that the presence of high levels of non-neutralizing antibodies in middle-aged adults has contributed to the continued persistence of 3c2.A viruses in the human population. Our findings might also relate to the unusual age distribution of H3N2 infections during the 2017-2018 season, in which H3N2 activity in middle-aged and older adults peaked earlier compared to children and young adults (9).

It is interesting that we detected significant NA antigenic change, but only minor HA antigenic change in viral isolates from the severe 2017-2018 season. We speculate that individuals with non-neutralizing HA antibody responses might rely on NA immunity, and it is possible that this has led to immune pressure on NA and the emergence of viruses with antigenically relevant NA substitutions. We found that NA substitutions in 3c2.A2 viruses from the 2017-2018 season reduced antibody recognition in serum from all age groups but especially from individuals 25 years old and younger. Further studies should be designed to tease out the protective role and interplay of HA and NA antibodies during the 2017-2018 season.

Our findings might shed light on the relatively low effectiveness of 3c2.A H3N2-based vaccines over the past 3 years (15–17). Low effectiveness of 3c2.A H3N2-based vaccines is partially due to the inherent difficulties of preparing egg-based 3c2.A H3N2 antigens (18). 3c2.A H3N2 viruses cannot grow in fertilized chicken eggs without first acquiring an amino acid substitution that abrogates a key glycosylation site in HA antigenic site B, and this egg-adaptation can dramatically alter antigenicity (8,19). Our study demonstrates that it is difficult to elicit neutralizing antibody responses against 3c2.A H3N2 viruses in some individuals even after natural infection with these viruses. Therefore, irrespective of issues related to 3c2.A H3N2-egg adaptations, it may be inherently difficult to design 3c2.A H3N2 vaccine antigens that are able to elicit neutralizing antibodies in humans that were exposed early in childhood with H3N2 viruses in the 1960s and 1970s.

Future studies should continue to evaluate the specificity of influenza virus antibodies in differently aged individuals. A better understanding of immunity within the population and within individuals will likely lead to improved models that are better able to predict the evolutionary trajectories of different influenza virus strains. Large serological studies and basic immunological studies may ultimately shed light on why influenza vaccines elicit variable responses in individuals with different immune histories, while identifying barriers that need to be overcome in order to design better vaccines that are able to elicit protective responses in all age groups.

## Materials and Methods

### Human serum samples

Serum samples from 140 children (1 to 17 years old) were collected at the Children’s Hospital of Philadelphia (CHOP) between May-August of 2017. Serum samples were originally collected from children for lead testing and leftover de-identified samples were then used for this study. Serum samples from 212 adults (18-90 years old) were collected via the Penn BioBank between May-August of 2017. The Penn BioBank routinely collects serum samples from individuals visiting the University of Pennsylvania Health care system. We did not include samples collected by the Penn BioBank from donors who had a pregnancy reported during the last 9 months, who had a medical history of cancer or organ transplantation, or who had a reported infectious disease within the previous 28 days. We also tested serum collected before and after laboratory confirmed 3c2.A H3N2 infections during the 2014-2015 and 2016-2017 influenza seasons. The 2014-2015 samples were collected from individuals participating in the Household Influenza Vaccine Effectiveness (HIVE) study in Ann Arbor, Michigan (14). Serum samples from this study were collected 0-3 months before and 4-8 months after laboratory-confirmed 3c2.A H3N2 infection. The 2016-2017 samples were collected from individuals infected in Rochester, NY. Samples from this study were collected at time of laboratory confirmed 3c2.A H3N2 infection (referred to in this manuscript as ‘pre-infection’) and 28 days later (post-infection samples). All studies involving human samples were approved by the Institutional Review Boards of the University of Michigan, University of Rochester, and the University of Pennsylvania.

### Viruses and recombinant proteins

All viruses used in this study were generated by reverse-genetics. HA and NA genes from A/Colorado/15/2014 (3c.2A) or A/Pennsylvania/49/2018 (3c.2A2) were cloned into the pHW2000 reverse genetics plasmid as described (20). Viruses were rescued using HA and NA genes from A/Colorado/15/2014 or A/Pennsylvania/49/2018 H3N2 viruses and internal genes from A/Puerto Rico/8/1934. For NA experiments, we generated viruses with H6 (A/turkey/Massachusetts/3740/1965) and N2 (from either A/Colorado/15/2014 or A/Pennsylvania/49/2018). Viruses were rescued by transfecting reverse-genetics plasmids into co-cultures of 293T and MDCK-SIAT1 cells. Transfection supernatants were harvested 3 days after transfection and stored at -80°C. HA and NA genes were sequenced to confirm that additional mutations did not arise during transfection. For the production of recombinant HA proteins, 1×10^8^ 293F cells were transfected with 70 μg codon-optimized A/Colorado/15/2014 HA plasmid in the presence of 10 μg NA plasmid using polyethylenimine transfection reagent. Proteins were purified after 3 days and the amount of recombinant HA for both constructs was normalized prior to use in ELISA.

### Foci Reduction Neutralization Test (FRNT)

Prior to testing in FRNT, serum samples were treated with receptor-destroying enzyme (RDE, Denka Seiken) for 2 hours at 37°C and the enzyme was then heat-inactivated at 55°C for 30 minutes. RDE-treated serum samples were serially diluted in twofold in MEM (Gibco). Virus was diluted to a concentration of approximately 300 focus-forming units per well and then incubated with serum samples for 1 hour at room temperature. Confluent monolayers of MDCK-SIAT1 cells were washed twice with MEM and then 100 μL of virus-serum mixtures were added to each well in a 96-well plate. Cells were incubated for 1 hour at 37°C in 5% CO_2_ and then washed with MEM. An overlay of 1.25% Avicel in MEM supplemented with 0.2% gentamicin and 1% 1M HEPES was then added to the cells. After 18 hours incubation at 37°C in 5% CO_2_, cells were fixed with 4% paraformaldehyde for 1 hour at 4°C. Cells were then lysed with 0.5% Triton X-100 in PBS for 7 minutes followed by blocking with 5% milk in PBS for 1 hour at room temperature. Plates were washed and then 50 μL of anti-NP monoclonal antibody IC5-1B7 (BEI) 1:5000 diluted in 5% milk in PBS was added to each well. After incubation for 1 hour at room temperature, plates were washed and 50 μL of anti-mouse peroxidase-conjugated secondary antibody (Fisher) 1:1000 diluted in 5% milk in PBS was added to each well. Plates were incubated for 1 hour at room temperature, washed, and then TMB substrate (Kirkegaard & Perry Laboratories) was added for visualization of the foci. Plates were imaged and foci were quantified using an ELISpot reader. FRNT_90_ titers are reported as the reciprocal of the highest dilution of sera that reduced the number of foci by at least 90%, relative to control wells that had no serum added. An anti-A/Colorado/15/2014 polyclonal serum in-house control was included in each assay run. Serum samples that did not reduce the number of foci by at least 90% reduction at a 1:20 serum dilution were assigned an FRNT_90_ titer of 10.

### Enzyme-Linked Immunosorbent Assay (ELISA)

ELISA plates were coated overnight at 4°C with recombinant HA protein. The next day, ELISA plates were blocked for 2 hours with PBS containing 3% BSA. Plates were washed 3 times with PBS containing 0.1% Tween 20 (PBS-T) and 50 μL of diluted serum was added to each well. After 2 hours of incubation, plates were washed 3 times with PBS-T and then 50 μL peroxidase-conjugated anti-human secondary antibody (Jackson ImmunoResearch) diluted in PBS containing 3% BSA was added to each well. After an hour incubation, plates were washed 3 times with PBS-T. SureBlue TMB substrate (KPL) was added to develop the plates, and 5 minutes later 250 mM hydrochloric acid was added to stop the reaction. Plates were read at an optical density (OD) of 450 nm using the SpectraMax 190 microplate reader (Molecular Devices). Monoclonal antibody 041-10047-1C04 was included on each plate as a control, and IgG concentrations were calculated based on the titration curve of this antibody (21). After background subtraction, serum samples that had an IgG concentration below the limit of detection were assigned an IgG concentration of 0.5 μg/mL.

### Enzyme-Linked Lectin Assay (ELLA)

ELLAs were completed to measure NA-specific antibody titers. Microtiter 96-well plates were coated with fetuin diluted in coating solution (KPL) and incubated overnight at 4°C. Heat-inactivated serum samples were serially diluted twofold in PBS containing 1% BSA and 0.1% Tween 20. Coated plates were washed with PBS-T and 50 μL of diluted serum was added to each well. Virus was diluted to the appropriate concentration and 50 μL of virus was added to each well. Plates were incubated overnight at 37°C. The next day, plates were washed with PBS-T and peanut agglutinin conjugated to HRP (PNA-HRP; Sigma) diluted in PBS containing 1% BSA was added to each well. Plates were incubated for 1 hour at room temperature and then washed with PBS-T. SureBlue TMB substrate (KPL) was added to each well to develop the plates and after 5 minutes the developing process was stopped with 250 mM hydrochloric acid. Plates were read at an optical density (OD) of 450 nm using the SpectraMax 190 microplate reader (Molecular Devices). After background subtraction, ELLA_50_ titers were reported as the reciprocal of the highest dilution of sera that reduced the OD value by at least 50%, relative to control wells that had no serum added. Serum samples that did not show at least 50% reduction in the OD value at a 1:20 serum dilution were assigned an ELLA_50_ titer of 10.

### Modeling

#### Imprinting probability

To calculate the probability of being imprinted with H3N2, we followed the approach of Gostic *et al*. 2016 (4) and estimated the risk of primary H3N2 infection for each birth cohort using surveillance data on the relative intensity of the influenza season, the percentage of influenza A isolates of each subtype that season, and typical attack rates in young children. We defined an influenza season *t* as from week 40 of year *t*-1 to week 39 of year *t*. The intensity of influenza A in season *t, I*_*t*_, was calculated as

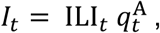

where ILI_*t*_ is the mean fraction of all people with influenza-like illness (ILI) in season *t*, and 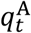 is the proportion of respiratory specimens testing positive for influenza A out of all respiratory specimens tested in season *t*. For the 1997-98 to the 2016-17 seasons, data on ILI and respiratory specimens were obtained from the U.S. Outpatient Influenza-like Illness Surveillance Network (ILINet) and World Health Organization/National Respiratory and Enteric Virus Surveillance System (WHO/NREVSS) Collaborating Labs (22). For the 1976-77 to the 1996-97 seasons, ILI was assumed to be the mean ILI of the above seasons, and the numbers of respiratory specimens were obtained from Thompson *et al*. 2003 (23). Each *I*_*t*_ from the 1976-77 season to the 2016-17 season was normalized by dividing by the mean of *I*_*t*_ across the seasons. For seasons from 1918-19 to 1975-76, we assumed that the intensity of influenza A to be 1.

We calculated the probability of having the first influenza A exposure to subtype H3N2 by taking the approach of Gostic *et al*. 2016 (4), assigning the probability of infection for naïve individuals a value of 0.28. Then the seasonal attack rate α was calculated by assuming exponential hazard as in Arevalo *et al*. 2019 (24):

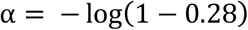

First, we calculated the probability of having the first influenza A exposure in each season for individuals of the same age. We defined individuals of age *a* to be born in time *y*, from the beginning of July in year *y*-1 to the end of June in year *y*, where year *y* is 2017 - *a*. The probability of having the first influenza A exposure in season *t* for individuals born in time *y* is

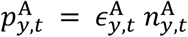

where 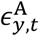 is the probability of infection with influenza A for naïve individuals born in time *y* in season *t*, and 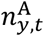 is the probability of being naïve to influenza A at the beginning of season *t*. Specifically,

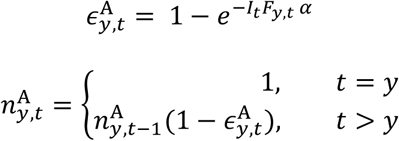

Here, *F*_*y,t*_, a fraction of a season *t* experienced by individuals born in time *y*, was introduced because individuals younger than 6 months of age are assumed not be exposed to influenza due to maternal immunity (see below).

Next, the probability that a cohort born in time *y* had its first influenza A exposure with subtype H3N2, *P*_*y*_, is calculated as

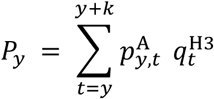

where 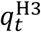 is the frequency of H3N2 among influenza A cases in season *t*, and *k* is the maximum age of having a primary influenza A infection (effectively a numerical correction). Here, we set *k* to be 20 for individuals of 20 years or older, and to be the age for individuals younger than 20 years. The choice of *k* minimally affected the value of *P*_*y*_.

#### Fraction of season experienced

We calculated the fraction of each season experienced by each cohort following the approach of Arevalo *et al*. 2019 (24). First, we calculated the fraction of a given influenza season during the week *w* of season *t, f*_*w,t*_ as

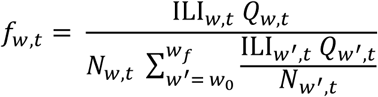

where ILI_*w,t*_ is the weighted fraction of patients with ILI in week *w* of season *t, Q*_*w,t*_ is the number of respiratory specimens testing positive to influenza A in week *w* of season *t*, and *N*_*w,t*_ is the number of specimens tested in week *w* of season *t*. Parameters *w*_0_ and *w*_*f*_ are the first and the final week of season *t*. For seasons before 1997-98, weekly data were unavailable. Therefore, for these seasons, we used the mean *f*_*w,t*_ of a given week *w* from the 1997-98 to the 2017-18 seasons. We calculated the fraction of an influenza season experienced by an individual born in time y, *F*_*y,t*_, using *f*_*w,t*_ and the proportion of individuals born in time *y* that are over 6 months old in week *w* of season *t, p*_*y,w,t*_:

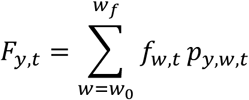

#### Amino acid similarity between the imprinting H3 virus and a test virus

To calculate the amino acid similarity between H3 viruses encountered early in life and a test virus, we calculated the probability of having the first influenza H3N2 exposure in season *t* for individuals born in time *y* as

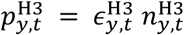

where 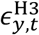 is the probability of H3N2 infection for naïve individuals born in time *y* in season *t*, and 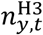 is the probability of being naïve at the start season *t*, i.e.,

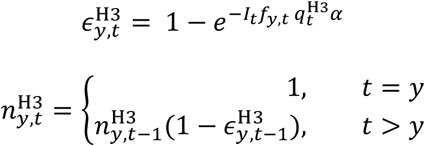

To calculate amino acid similarity, the H3 sequence samples were downloaded from Global Initiative on Sharing All Influenza Data in August 2018 (25). For each season, we resampled 100 samples if the number of available sequences within the season exceeded 100. The amino acid similarity between H3N2 strains circulating in season *t* and a test virus *v, s*_*t,v*_, was calculated as the fraction of shared amino acids in HA head epitopes (26),

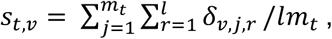

where *m*_*t*_ is the number of H3 sequence samples collected in season *t, j* is the index for the sample, *l* is the number of residues in the H3 head epitopes, *r* is the index for the residue, and *δ*_*j,r,v*_ is an indicator for the match of the amino acid between a test virus *v* and sample *j* at residue *r*. The value of *δ*_*j,r,v*_ is 1 if the amino acids match and 0 otherwise. For seasons before 1991-92, where the precise date of sequence isolation was unavailable, we obtained *s*_*t,v*_ by calculating the average of *s*_*v*_ of year *t*-1 and year *t*, weighted by the average fraction of sequences from year *t*-1 and from year *t* for season *t*, calculated from seasons from 1991-92 to 2016-17.

Finally, the expected amino acid similarity between the first H3 virus encountered in life and a test virus *v* for individuals born in time *y* is

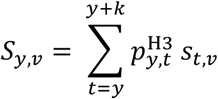

We assumed that individuals born before 1967 to have the same amino acid similarity as individuals born in 1967.

#### Regression model

To investigate associations between H3 imprinting, the amino acid similarity between the first H3 virus encountered in life and a test virus, and titers to the test virus, we performed a linear regression,

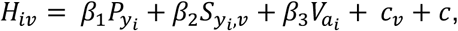

where *H*_*i,v*_is individual *i*’s log_2_ FRNT titer to test virus *v*, 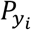 is the probability of imprinting to H3 for individual *i* born in birth time *y*, 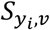 is the expected amino acid similarity between the first H3 virus encountered by the birth cohort to which individual *i* belongs and test virus *v*, 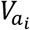 is the vaccination coverage for people the same age as individual *i, c*_*v*_ is a virus-specific intercept, and *c* is constant. Age-specific vaccine coverage from Pennsylvania state for the 2016-17 season was obtained from National Immunization Survey-Flu (NIS-Flu) and Behavioral Risk Factor Surveillance System (BRFSS) (26). We also evaluated models with a linear age effect as

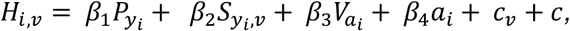

where *β*_4_ is the coefficient for age. Finally, we evaluated whether there was support for titer differences in two epidemiologically distinct age groups, children (<18 years old) and adults (≥ 18 years old):

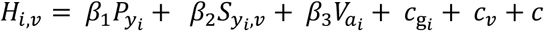

where 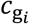 is the effect for age group *g* of individual *i*. We used the glm function in R (version 3.4.2) to fit linear models.

### Statistical analysis

Data are presented as geometric mean antibody levels with 95% confidence intervals (CI) if not stated otherwise. Analyses were performed using GraphPad Prism (version 7) or R (version 3.4.2). To compare titers between age groups, we performed bootstrap tests according to Hall and Wilson 1991 (27). For a given pair of age groups, we calculate mean titer difference, 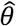, and tested if it is significantly greater than zero. The bootstrap value of 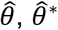, is obtained by resampling individuals in each age group with replacement. We obtained the distribution of 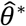 from 20,000 resamplings. The distribution of 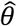 under a null hypothesis of 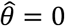 is 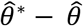 and we calculated p values from this null distribution. Bootstrapping was performed for every pair of age groups. We then used Bonferroni correction for multiple comparisons by dividing 0.05 by 72, the number of all pairs of age groups. Therefore, a null hypothesis is rejected if p < 0.0007. Pre- and post-infection titers during the 2014-2015 season were log^2^-transformed and then compared using the paired t-test. P values <0.05 were considered as statistically significant.

## Data Availability

All data are in the manuscript. Raw data can be obtained from senior author after publication in peer reviewed journal.

## Acknowledgements

This work was supported by the National Institute of Allergy and Infectious Diseases (1R01AI113047, S.E.H.; 1R01AI108686, S.E.H.; 1R01AI097150, A.S.M.; CEIRS HHSN272201400005C, S.E.H., S.C., E.T.M., A.S.M. A.B., D.J.T.) and Center for Disease Control (U01IP000474, A.S.M.). Scott E. Hensley holds an Investigators in the Pathogenesis of Infectious Disease Awards from the Burroughs Wellcome Fund.

## Author contributions

S.G., M.W., and M.G. completed experiments and analyzed data. K.K. and S.C. analyzed data and completed modeling studies. A.B., D.J.T., E.T.M., and A.S.M. provided serum samples from individuals before and after PCR-confirmed H3N2 infection. S.E.H. conceived the project and supervised the work. S.G. and S.E.H. wrote the manuscript with input from all authors.

## Competing interests

S.E.H. has received consultancy fee from Sanofi Pasteur, Lumen, Novavax, and Merck for work unrelated to this report. A.S.M. has received consultancy fees from Sanofi Pasteur, Seqirus, and Novavax for work unrelated to this report. A.B. reports has received consultancy fees from GSK and Merck and grant support from Merck, Pfizer, and Janssen for work unrelated to this report. All other authors report no potential conflicts.

**Extended Data Table 1.**
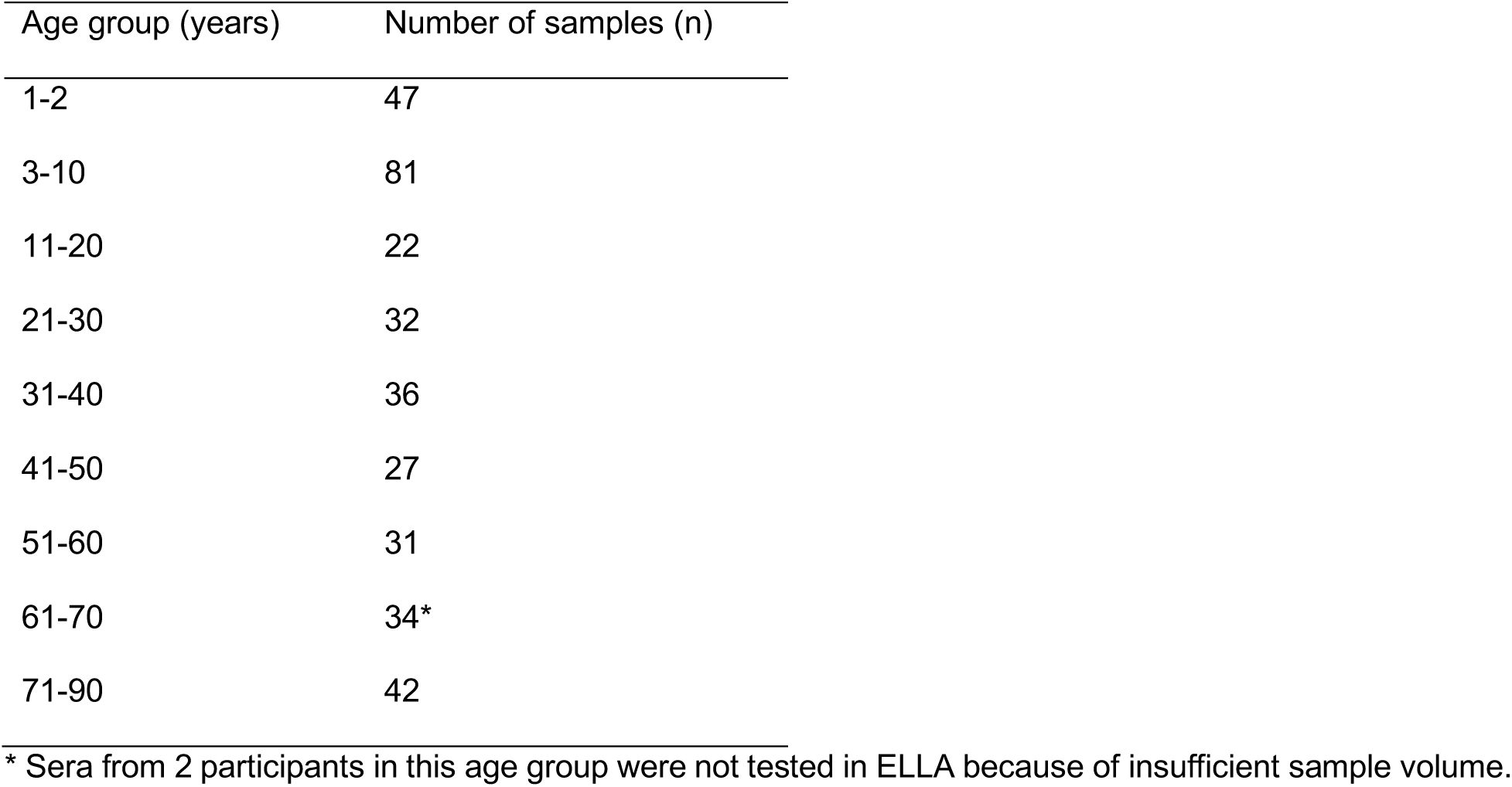
Number of serum samples per age group.

**Extended Data Table 2.**
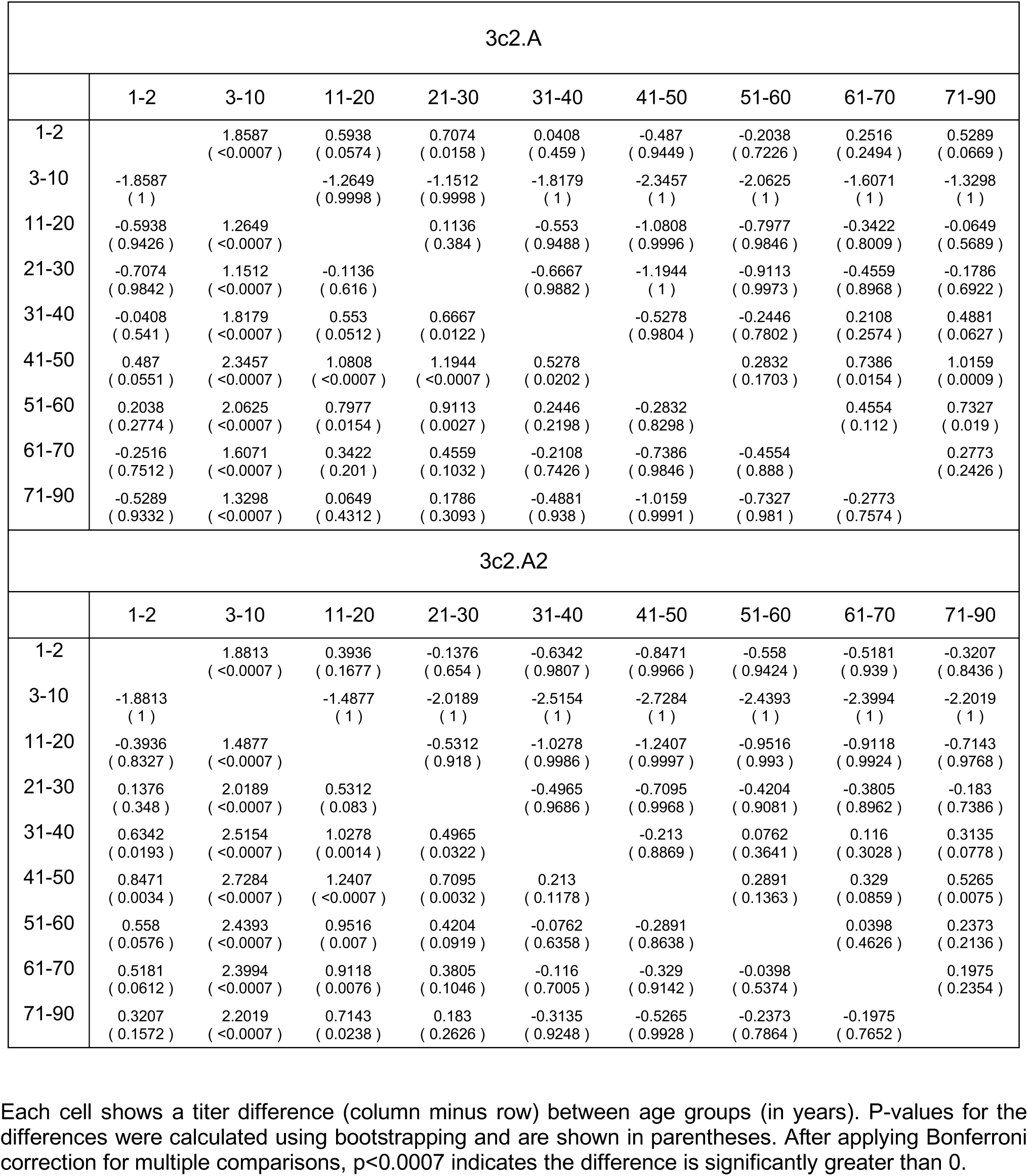
Comparisons of FRNT titers between age groups.

**Extended Data Table 3.** The estimates of the effects of imprinting, vaccine coverage effect, and virus specific effect.

| Variable | Estimate (standard error) | p | AIC |
| --- | --- | --- | --- |
| Model 1 |  |  | 2647.5 |
| Imprinting to H3 | 0.17 (0.30) | 0.58 |  |
| Amino acid similarity between first H3 virus in life and a test virus | 3.45 (0.38) | <0.001 |  |
| Vaccine coverage | 3.86(0.56) | <0.001 |  |
| Virus effect for 3c2.A2 | -0.23(0.12) | 0.05 |  |
| Model 2 (with linear age effect) |  |  | 2648.9 |
| Imprinting to H3 | 0.33 (0.36) | 0.37 |  |
| Amino acid similarity between first H3 virus in life and a test virus | 3.57 (0.41) | <0.001 |  |
| Age | 0.003 (0.003) | 0.44 |  |
| Vaccine coverage | 4.19 (0.71) | <0.001 |  |
| Virus effect for 3c2.A2 | -0.23 (0.12) | 0.06 |  |
| Model 3 (with age group effect for <18) |  |  | 2649.4 |
| Imprinting to H3 | 0.12 (0.32) | 0.71 |  |
| Amino acid similarity between first H3 virus in life and a test virus | 3.37 (0.44) | <0.001 |  |
| Age group <18 | 0.08 (0.24) | 0.73 |  |
| Vaccine coverage | 3.60 (0.94) | <0.001 |  |
| Virus effect for 3c2.A2 | -0.23 (0.12) | 0.05 |  |

**Extended Data Figure 1.**
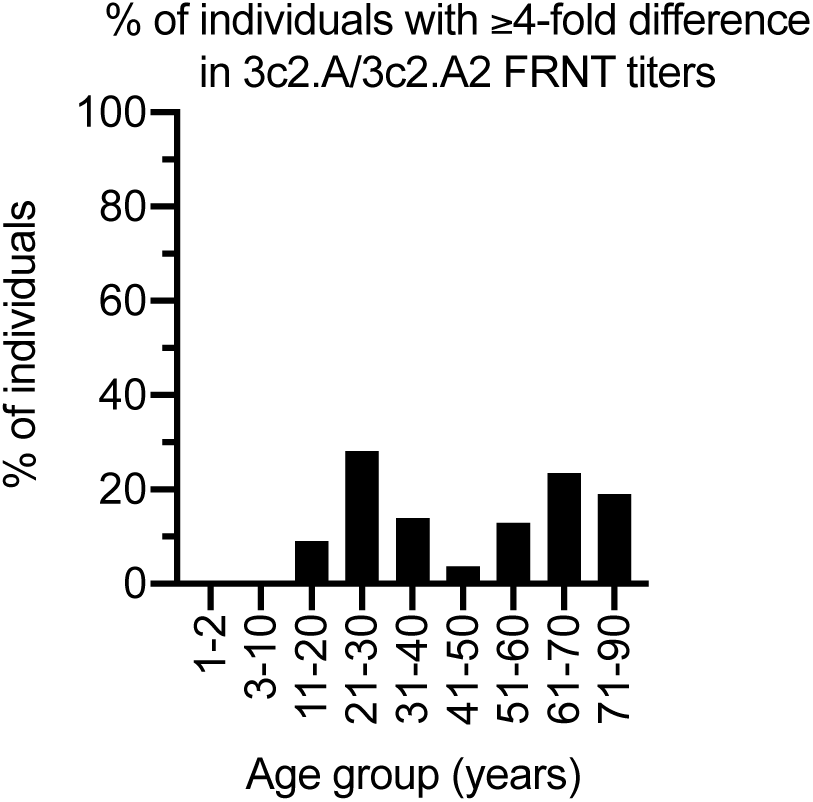
Percentage of individuals with a ≥4-difference in neutralizing antibody titers as measured in FRNT in Figure 1.

**Extended Data Figure 2.**
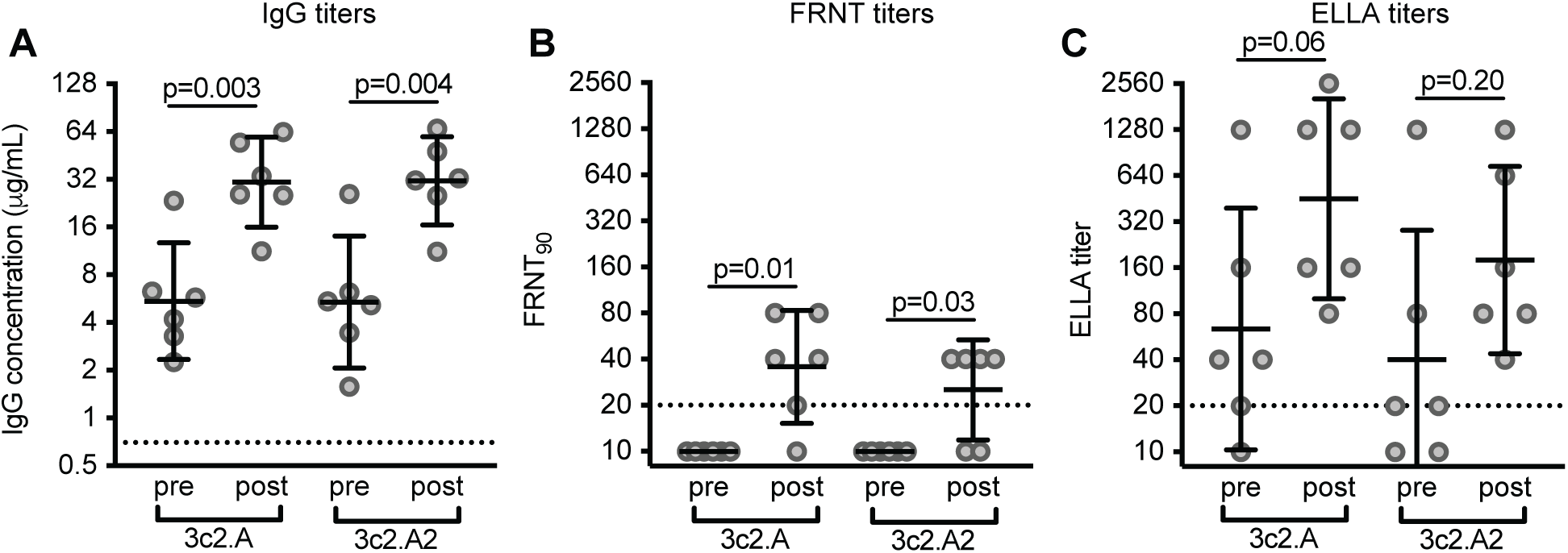
Antibody responses in adults before and after laboratory-confirmed 3c2.A H3N2 infection during the 2016-2017 influenza season. Serum was collected from 6 adults before and after laboratory-confirmed 3c2.A H3N2 infection and we quantified (A) HA IgG binding antibodies by ELISA, (B) neutralizing antibodies by FRNTs, and (C) NA-specific antibodies by ELLAs. Dashed lines represent the limit of detection for each assay. Lines represent the geometric mean titers with 95% confidence intervals. Paired t tests were completed on log_2_-transformed data, and p values are indicated.

